# From preconception to postpartum: bidirectional associations between sleep and depression and the role of infant sleep in a population-based cohort

**DOI:** 10.64898/2026.08.26.26361397

**Authors:** Fangxiang Mao, Hanan EI Marroun, Sanne J.W. Hoepel, Susan J. Ravensbergen, Isabel K Schuurmans

**Affiliations:** The Generation R Study Group, Erasmus MC University Medical Center, Rotterdam, Zuid-Holland, the Netherlands; Department of Child and Adolescent Psychiatry/Psychology, Erasmus MC University Medical Center, Rotterdam, Zuid-Holland, the Netherlands; Department of Psychology, Education and Child Studies - School of Social and Behavioural Sciences, Erasmus University Rotterdam, Rotterdam, Zuid-Holland, the Netherlands; Department of Epidemiology, Erasmus MC University Medical Center, Rotterdam, Zuid-Holland, the Netherlands; Department of Psychiatry, Erasmus MC University Medical Center, Rotterdam, Zuid-Holland, the Netherlands

**Author notes:** Corresponding author: Hanan EI Marroun, Academic affiliation: Department of Child and Adolescent Psychiatry/Psychology, Erasmus MC University Medical Center, Rotterdam, Zuid-Holland, the Netherlands. Department of Psychology, Education and Child Studies - School of Social and Behavioural Sciences, Erasmus University Rotterdam, Rotterdam, Zuid-Holland, the Netherlands. Mail Address: Dr. Molewaterplein 40, 3015 GD, Rotterdam, Zuid-Holland, the Netherlands.

**Keywords:** sleep, depressive symptoms, perinatal, preconception, population-based, cohort study

## Abstract

This study investigated bidirectional associations between maternal sleep and depressive symptoms from preconception to postpartum, and whether infant sleep mediated or moderated these associations. We used data from the Generation R *Next* Study (N=2,294). Maternal sleep (specifically general sleep disturbance, latency, quality, duration, and midpoint) and depressive symptoms were prospectively assessed at five timepoints from preconception to 12-month postpartum. Sleep was self-assessed with the General Sleep Disturbance Scale and Munich Chronotype Questionnaire; depressive symptoms with the Adult Self Report depression/anxiety subscale and Edinburgh Postnatal Depression Scale. Infant sleep (specifically night awakenings, nocturnal sleep duration, and latency) was parent-reported at 1-month postpartum using the Brief Infant Sleep Questionnaire. Bidirectional associations were examined using Autoregressive Latent Trajectory Models with Structured Residuals. The role of infant sleep was examined using mediation and moderation analyses. We found that maternal sleep and depressive symptoms were both stable over time. For sleep quality and disturbance, bidirectional associations suggested slightly stronger effects from depression to sleep (sleep quality:β_depressionàsleep quality_=0.11, 95%CI:0.07–0.14; general sleep disturbance:β_depressionàsleep disturbance_=0.14, 95%CI:0.10–0.18) than from sleep to depression (β_sleep quality/disturbanceàdepression_=0.07 for both, 95%CIs:0.03–0.11). For latency, effects were comparable in both directions (β_depressionàsleep latency_=0.06, 95%CI:0.03–0.09; β_sleep latencyàdepression_=0.05, 95%CI:0.01–0.09). The association between depressive symptoms and sleep latency was both mediated (9.7%) and moderated (p<0.05) by infant sleep latency. In conclusion, general maternal sleep disturbance, sleep quality, and sleep latency showed bidirectional associations with depressive symptoms from preconception/early pregnancy onwards. Infant sleep latency may represent a potential modifiable factor within this cycle.

## Introduction

Sleep and depressive symptoms are closely associated across the life course (Pandi-Perumal et al., 2020). Evidence from both general and clinical populations indicates this relation is bidirectional: sleep problems can be considered a prodromal symptom of depression (American Psychiatric Association, 2013), while depressive symptoms can, in turn, also predict subsequent sleep problems (American Psychiatric Association, 2013; Mookerjee et al.; Singareddy et al.; Yao et al.). Longitudinal studies further demonstrate that sleep and depressive symptoms can exacerbate one another over time (Bouwmans et al., 2017; Cousins et al., 2011; de Feijter et al., 2023; Jansson-Fröjmark and Lindblom, 2008; Saunders et al., 2023; Sivertsen et al., 2012; Sun et al., 2018). However, while this bidirectional association is well-established across the life course, its dynamics remain under-investigated across the perinatal period, a phase characterized by profound physical and psychosocial changes that predispose women to both poor sleep and depressive symptoms (Silvestri and Aricò, 2019; The, 2023). Clarifying the bidirectional association between sleep and depression from preconception to postpartum is crucial to inform targeted interventions, helping to determine whether prioritizing perinatal sleep, depressive symptoms, or both may prevent and redirect this cycle.

Existing studies on depressive symptoms and sleep across the perinatal period are mostly cross-sectional, focusing on a single timepoint without repeated measurements (Emamian et al., 2019; Fu et al., 2023; Lawson et al., 2015; Li et al., 2023; Maghami et al., 2021; Sobol et al., 2023). The few studies that adopted prospective designs almost exclusively investigated prenatal sleep as an exposure and postnatal depressive symptoms as the outcome, showing that poor sleep predicts subsequent depressive symptoms (Li et al., 2023). This temporal direction from sleep to depressive symptoms is further supported by randomized controlled trials showing that improving prenatal sleep quality can reduce the risk of postnatal depressive symptoms (Quin et al.; Silang et al.). Furthermore, existing evidence has primarily focused on associations between sleep and depressive symptoms from the third trimester to three months postpartum (Li et al., 2023), overlooking both earlier stages, such as preconception–a sensitive window for maternal and child health (Bais et al., 2022)–and later stages up to 12 months postpartum. Consequently, little is known about the full bidirectional dynamics of sleep– depressive symptom association across the entire perinatal period (from preconception to postpartum).

Notably, during the postpartum period, infant sleep is introduced as a third factor that may further shape the bidirectional association between perinatal sleep and depressive symptoms. Poor prenatal maternal sleep has been associated with greater infant sleep problems (e.g., late bedtimes, frequent awakenings, short nighttime sleep) (Ciciolla et al., 2022; Hoyniak et al., 2024; Lahti-Pulkkinen et al., 2019; Nakahara et al., 2020), which in turn are associated with elevated maternal depressive symptoms (Dias and Figueiredo, 2021; Gui et al., 2022; Petzoldt et al., 2016; Škodová et al., 2022; Warren et al., 2006). Randomized controlled trials provide further support by showing that infant sleep education can significantly reduce maternal sleep problems (Rouzafzoon et al., 2021) or depressive symptoms (Hiscock et al., 2014). A cross-sectional study that investigated maternal depressive symptoms and maternal and infant sleep simultaneously found more severe postnatal depressive symptoms in mothers whose prenatal sleep duration was misaligned with their infant’s sleep patterns (Newland et al., 2016). While these findings suggest that infant sleep acts as a mediator or moderator in the association between maternal sleep and depressive symptoms, this has rarely been examined within the same longitudinal study.

Therefore, this study aimed to (1) explore the bidirectional longitudinal associations of maternal sleep and depressive symptoms starting from preconception through pregnancy to 12 months postpartum in a population-based cohort, and (2) examine whether infant sleep mediated or moderated these associations. Recognizing sleep as a multidimensional construct (Buysse, 2014), we assessed multiple sleep characteristics separately for both mothers and infants, including in mothers, sleep duration, sleep midpoint (the clock time midway between sleep onset and offset), sleep latency (time to fall asleep), sleep quality (self-rated quality of sleep experience), and general sleep disturbance, and in infants, nocturnal sleep duration, number of nighttime awakenings, and sleep latency. The conceptual framework is presented in Figure 1.

**Figure 1:**
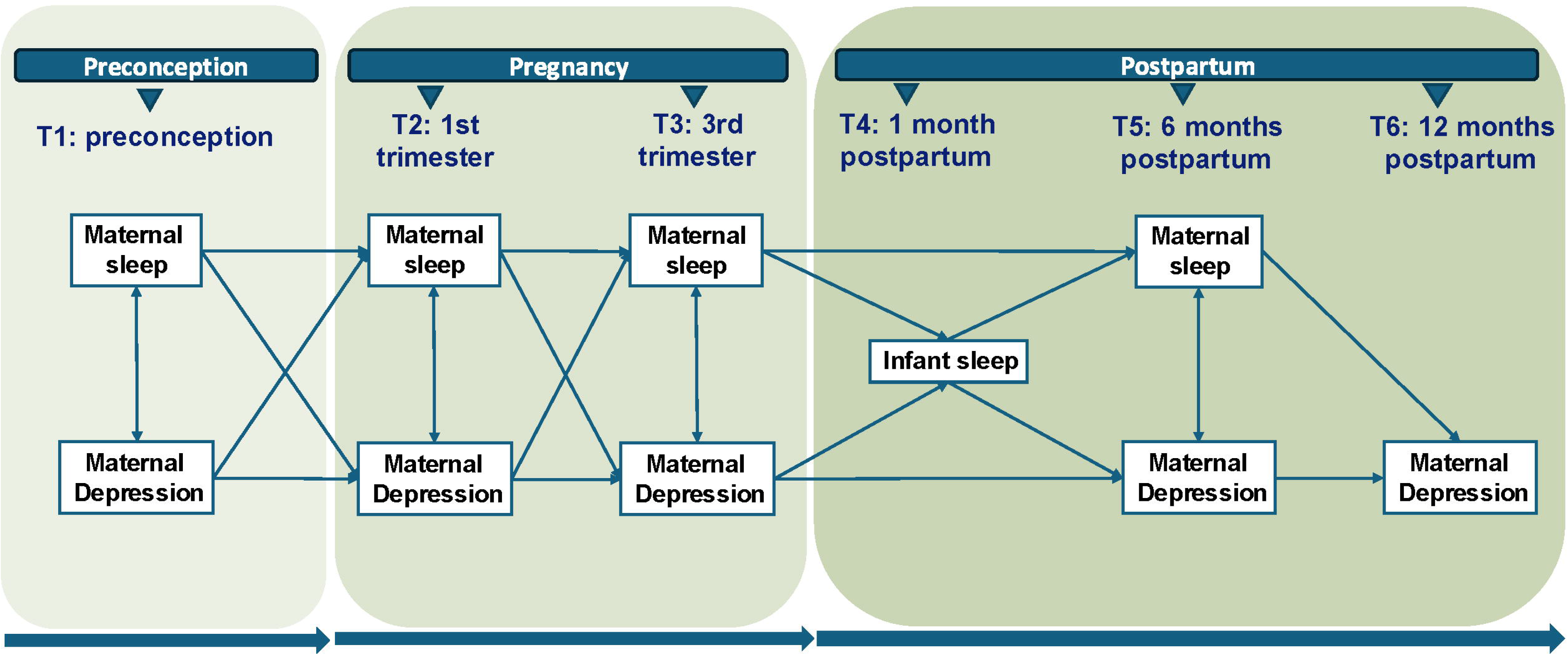
Framework of this study. *Figure notes:* Covariates were adjusted in the model, including maternal age at enrollment, country of birth, educational status, household net income monthly, living with a partner [yes/no], parity before enrollment, body mass index at enrollment [kg/m^2^], smoking history in the past 12 months, alcohol use history in the past 12 months, and gestational age at each time point. Given the moderate to high autocorrelations of sleep and depressive symptoms over time (see heatmap in Figure S2), the weights of all autoregressive paths (stability over time) and all cross-lagged paths (directionality over time) were separately constrained to be equal in the main analysis

## Methods

### Participants

This study is embedded in the Generation R *Next* Study, a population-based prospective cohort in Rotterdam, the Netherlands (Boxem et al., 2024). The aim of the Generation R *Next* Study is to identify preconception and early-pregnancy determinants of fertility, embryonic development, and childhood outcomes. The study procedures were approved by the Medical Ethical Committee of the Erasmus Medical Center (MEC-2016-589, December 2016). Written informed consent was obtained from all participants.

Women were eligible if (1) they were aged 18 years or older, (2) lived in Rotterdam (zip code 3011-3099 & 3191-3196), and (3) were planning to become pregnant (preconception period) or were pregnant and had not yet given birth. The inclusion period spanned between August 9, 2017, and July 1, 2021. Using study questionnaires and visits at the research center, data was repeatedly collected from the time of inclusion i.e., at preconception, first trimester, third trimester, six months postpartum, and 12 months postpartum. Women could participate in the study multiple times. Each participation could result in a singleton or twin birth. A total of 4,082 participants were enrolled.

Of the 4,082 participants, we excluded those without postpartum follow-up (n = 1,140, Figure 2). In case the participant took part in the study multiple times or with twins, only one enrollment per participant was retained, and additional enrollments were randomly excluded (n = 179), to ensure independence among participants. We further excluded women with no data on sleep (n = 441) or depressive symptoms (n = 28). The final study sample consisted of 2,294 unique participants with data available for at least one of four sleep assessments and one of five depressive symptom assessments. Supplementary Figure S1 visualizes the frequency of different combinations of available data over time.

**Figure 2:**
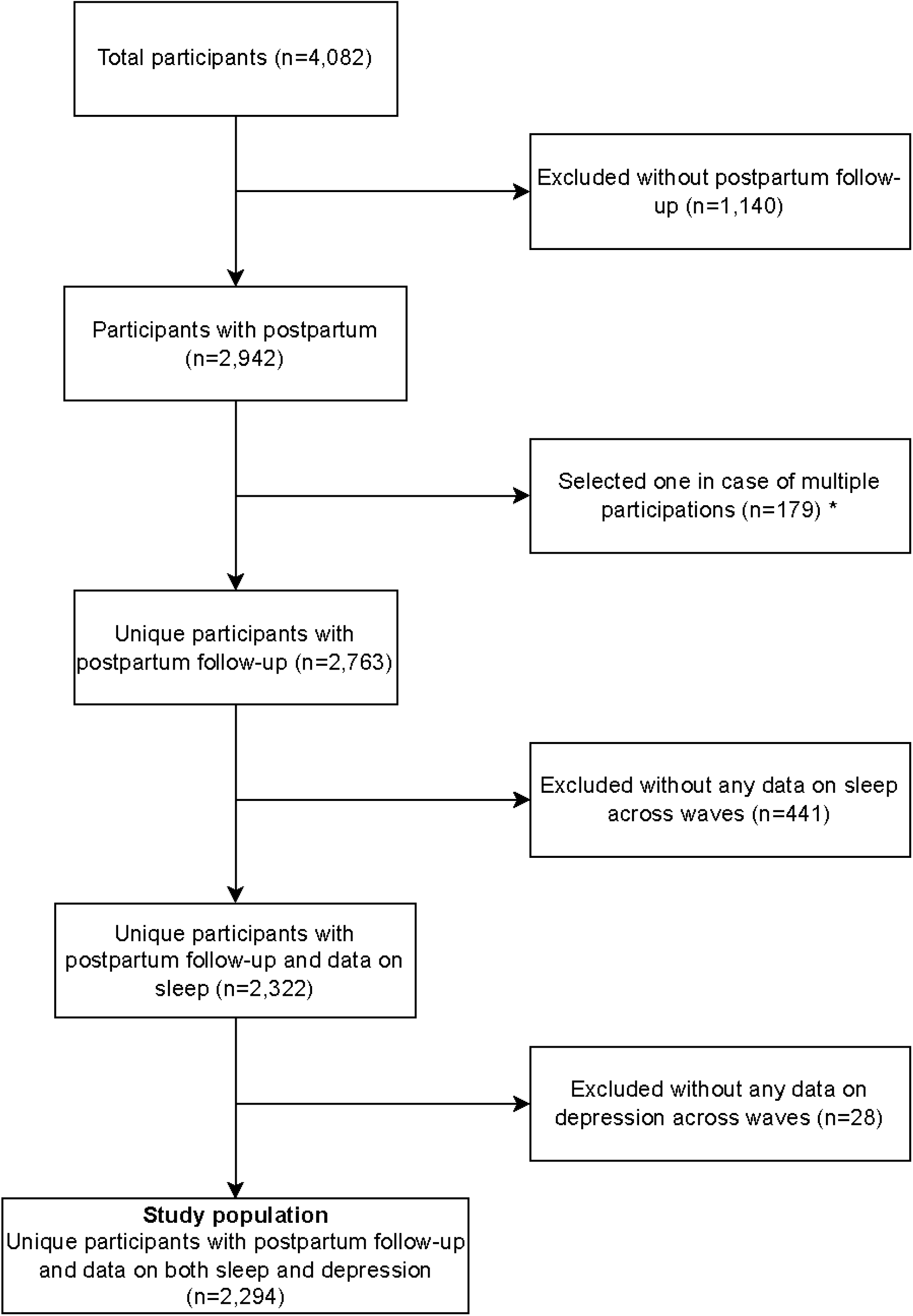
Flowchart of the study population. *Figure notes:* Women could participate in the study multiple times. In case of multiple participation, only one enrollment per woman was retained, and additional enrollments were randomly excluded (n = 44 for twins, n = 135 for multiple enrollments) to ensure independence among participants

### Measures

#### Maternal sleep

Maternal sleep was measured using two validated self-reported instruments: the Munich Chronotype Questionnaire (MCTQ) and the General Sleep Disturbance Scale (GSDS). Both were administered in parallel at four timepoints: preconception, first trimester, 3^rd^ trimester, and six months postpartum. The characteristics of sleep over time have been described in our previous study (Mao et al., 2025).

The MCTQ was used to assess sleep duration, latency and midpoint (Roenneberg et al., 2003). The Dutch version of MCTQ performs well in validity and reliability (Zavada et al., 2005). In this study, we used a shortened version with 12 items that assessed bedtime, intended time to fall asleep, sleep latency, wake up time, time until getting out of bed, and alarm use, separately for workdays and work-free days. Sleep data on work and work-free days were combined into one item:

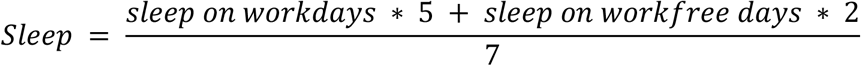

Based on all 12 items, we calculated sleep components as follows:

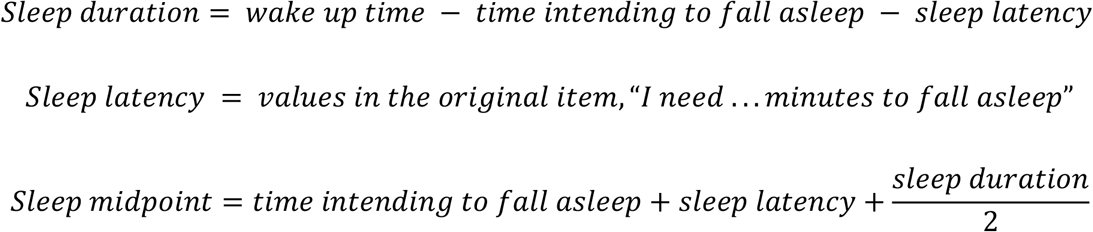

The GSDS was used to assess subjective sleep quality and general sleep disturbance (Lee and DeJoseph, 1992). The GSDS consists of 21 items and covers seven subdomains (sleep quality, sleep quantity, difficulty in initiating sleep, midsleep awakenings, early awakenings, medications for sleep, excessive daytime sleepiness) and one composite index (summed score of all items). Participants reported how many days in the past week they experienced these symptoms. Higher sum scores indicated poorer sleep. We included the subdomain sleep quality (range 0-21) and the overall score (general sleep disturbance, range 0-147) in our analyses. The GSDS has demonstrated validity in pregnant women (Coo et al., 2014). If missing items exceeded 25%, the total score was considered missing. If missing items was less than 25%, weighted sum scores (considering the number of endorsed items) were calculated.

#### Maternal depressive symptoms

Maternal depressive symptoms were assessed using two validated instruments administered at different stages of the study (not in parallel): the Adult Self-Report (ASR) Anxious/Depressed subscale and the Edinburgh Postnatal Depression Scale (EPDS). Both instruments are widely used in perinatal research and primarily capture depressive symptoms, although they also include items reflecting anxiety (Achenbach and Rescorla, 2003; Cox et al., 1987). This overlap is consistent with the high comorbidity of depression and anxiety during the perinatal period and supports their use as indicators of maternal depressive symptomatology (Achenbach and Rescorla, 2003; Matsumura et al., 2020; Wisner et al., 2013). Using both measures at different stages allowed us to capture depressive features relevant to both general and perinatal populations.

The ASR Anxious/Depressed subscale was assessed at preconception, the first trimester, and 12 months postpartum (Achenbach and Rescorla, 2003). The ASR includes 129 items, of which the empirically-derived Anxious/Depressed subscale contains 18 items (e.g., *“I feel worthless or inferior,” “I feel that no one loves me”*). Each item was rated from 0 (“Not true”) to 2 (“Very true or often true”), with higher summed scores reflecting greater depressive and anxiety symptoms. We applied the empirically derived syndrome-based symptom scoring rather than DSM-oriented scoring for two main reasons: first, our focus on a general population sample makes capturing the continuum of symptom severity more appropriate than diagnostic categories; and second, this approach enhances comparability with the EPDS, which was explicitly designed for perinatal populations by down-weighting somatic symptoms and emphasizing emotional and cognitive features, including anxiety (Cox et al., 1987). The validity of the ASR Anxious/Depressed subscale has been demonstrated in both population-based and clinical settings (Ivanova et al., 2015).

The EPDS was administered at the third trimester and six months postpartum. The EPDS is a widely used 10-item self-report scale that is sensitive to perinatal-specific symptoms (Cox et al., 1987; Pop et al., 1992). The EPDS consists of 10 items, and each item was rated from 0 (“No, never”) to 3 (“Yes, very often”). A higher total EPDS score reflects more symptoms of depression. Its reliability and validity have been demonstrated in Dutch women at postpartum (Pop et al., 1992).

To improve comparability between ASR and EPDS, raw scores were standardized within each timepoint before modelling. When item-level missing data were present, we applied the weighting method considering the number of endorsed items if missing <25% and >25% as missing.

#### Infant sleep

Infant sleep was measured only at one month postpartum, using three domains from the parent-reported Brief Infant Sleep Questionnaire (BISQ) (Sadeh, 2004). The BISQ originally includes 10 domains, including nocturnal sleep duration, daytime sleep duration, the number of night awakenings, nocturnal bedtime, wakeup time, sleep latency, routine or ritual for bedtime, location of falling sleep, location of sleep, and parents’ perception of infant sleep problem. In this study, we only included three of these domains: nocturnal sleep duration (self-reported hours of sleep between 7 pm and 7 am), the number of night awakenings (rated from 0 (“no awakenings”) to 7 (“7 times awakenings or more”)), and sleep latency (time to falling asleep for the night, self-reported duration). Other BISQ domains were not included because they are less reliable at one month of age (e.g., bedtime), reflect parental practices rather than infant sleep itself (e.g., bedtime routines), or are strongly influenced by parental perceptions and maternal mood (e.g., perceived sleep problems). Including these domains could have introduced measurement bias, reduced construct validity, and increased common method bias in relation to maternal depressive symptoms. The reliability and validity of the BISQ have been demonstrated in the Dutch population (Harskamp-van Ginkel et al., 2023).

#### Covariates

Confounding factors were selected based on literature (Chen et al., 2019; Dias and Figueiredo; Pauley et al., 2020; Qiu et al., 2022). Maternal sociodemographic factors were assessed by questionnaire at enrolment and included (Pauley et al., 2020): maternal age (in years); country of birth (within the Netherlands vs. outside the Netherlands); educational status (primary or secondary/vocational, bachelor’s or associate degree, master’s degree); household net monthly income (<€3000 vs. ≥€3000); living with a partner (*yes*: married and cohabiting, cohabiting but not married, or registered partnership and cohabiting vs. *no/other*: no partner or not cohabiting, widow, married but living separately, divorced/separated, or other); and parity (nulliparous vs. primiparous or multiparous). Also, self-reported lifestyle factors at enrollment were considered, including body mass index (BMI, kg/m^2^) (Pauley et al., 2020), lifetime smoking (never vs. ever or current) (Chen et al., 2019) and alcohol use in the past 12 months (yes vs. no) (Qiu et al., 2022). Further, we adjusted for gestational age at each time point of data collection, operationalized as the number of weeks from conception, with conception defined as time zero and preconception periods represented by negative values. Child sex (male vs. female) was obtained from birth records and included for descriptive purposes only (Dias and Figueiredo, 2020).

### Statistical analysis

We summarized categorical and continuous variables as counts (percentages) and means (standard deviations), respectively. We compared demographic differences between women included and excluded using independent-samples t-test, chi-squared test, or Fisher’s exact test.

To examine the bidirectional associations between each sleep component and depressive symptoms, we applied the Autoregressive Latent Trajectory Model with Structured Residuals (ALT-SR) using the R package *lavaan* (Muthén and Muthén, 2015). This model distinguishes changes within the same individual over time from differences between individuals, helping to avoid the bias that can occur when these effects are mixed together in traditional cross-lagged panel models (Berry and Willoughby, 2017). Missing data were handled using full information maximum likelihood. Model fit was evaluated using comparative fit index (CFI) and the root mean square error of approximation (RMSEA) (McDonald and Ho, 2002). A CFI value of ≥0.90 or an RMSEA value ≤0.06 was considered indicators of good fit (Ullman and Bentler, 2003). Given the moderate to high autocorrelations of sleep and depressive symptoms over time (Supplementary Figure S2), the weights of all autoregressive paths (stability over time) and all cross-lagged paths (directionality over time) were separately constrained to be equal in the main analysis. We assessed the mediating and moderating roles of infant sleep within the structural equation model framework, using 1,000 bootstrap iterations implemented with *lavaan*, limited to associations where significant bidirectional and autoregressive paths were identified (significance level p<0.05).

In the sensitivity analysis, we tested the robustness of results by replicating the ALT-SR model by (1) allowing all path weights to vary given the possibility that the strength of the sleep-depressive symptoms associations may change over time and (2) recalculating EPDS scores after removing item 7 (sleep difficulties) to reduce the influence of potential measurement overlap.

All analyses were conducted in R version 4.5.1. Two-sided tests with p<0.05 were considered statistically significant.

## Results

### Characteristics of the study population

Table 1 shows the descriptive statistics of the study population. Women included in the study were on average 31.7 ± 4.2 years old at enrollment. Two-thirds (63.6%) of women were born in the Netherlands. About 42.1% had a master’s degree. Most women (87.2%) lived with partners and were primiparous (65.8%). About half (44.5%) had ever smoked and three quarters (78.1%) had ever drunk alcohol.

**Table 1:** Characteristics of women included in the study.

| Demographic variables | Study population (n=2,294), Mean<br>± SD / No. (%) |
| --- | --- |
| Age at enrollment | 31.7 ± 4.2 |
| Country of birth |  |
| Within the Netherlands | 1423 (63.6%) |
| Outside the Netherlands | 814 (36.4%) |
| Education status |  |
| Primary or secondary/vocational | 603 (27.2%) |
| Bachelor's or associate degree | 681 (30.7%) |
| Master's degree | 933 (42.1%) |
| Household net income, monthly |  |
| Less than €3000 | 458 (22.4%) |
| Equal or more than €3000 | 1583 (77.6%) |
| Living with partner |  |
| Yes | 1903 (87.2%) |
| No or Other | 280 (12.8%) |
| Parity before enrollment |  |
| Nulliparous | 1450 (65.8%) |
| Primiparous or multiparous | 753 (34.2%) |
| Body mass index at enrollment, kg/m<sup>2</sup> | 24.2 ± 4.4 |
| Smoking status lifetime |  |
| Never | 1164 (55.5%) |
| Any | 932 (44.5%) |
| Alcohol use status lifetime |  |
| Never | 489 (21.9%) |
| Ever or current | 1746 (78.1%) |
| Child sex |  |
| Male | 1188 (52.7%) |
| Female | 1068 (47.3%) |

*Table Note:* The percentages of missing values across all variables ranged from 0.1 to 11.0%. The highest percentage of missing was found in maternal net family income monthly.

Compared to women excluded, women included in the analysis were older, were more often born in the Netherlands than outside the Netherlands, had a higher educational status, higher net family income monthly, were more often living with a partner than not living with a partner, were more often nulliparous, had a lower BMI, were more likely to not smoke, and were more likely to have ever used alcohol before inclusion in the study (Table S1).

### The association between sleep and depressive symptoms

Estimates are summarized in Figure 3, presenting the stability within domains over time (sleep↔sleep, depressive symptoms↔depressive symptoms, right panel) and directionality between sleep and depressive symptoms over time (sleepàdepressive symptoms, depressive symptomsàsleep, left panel). The bidirectional model showed good fit (CFI>0.75 and RMSEA <0.06). Detailed information of estimates is presented in Table S2.

**Figure 3:**
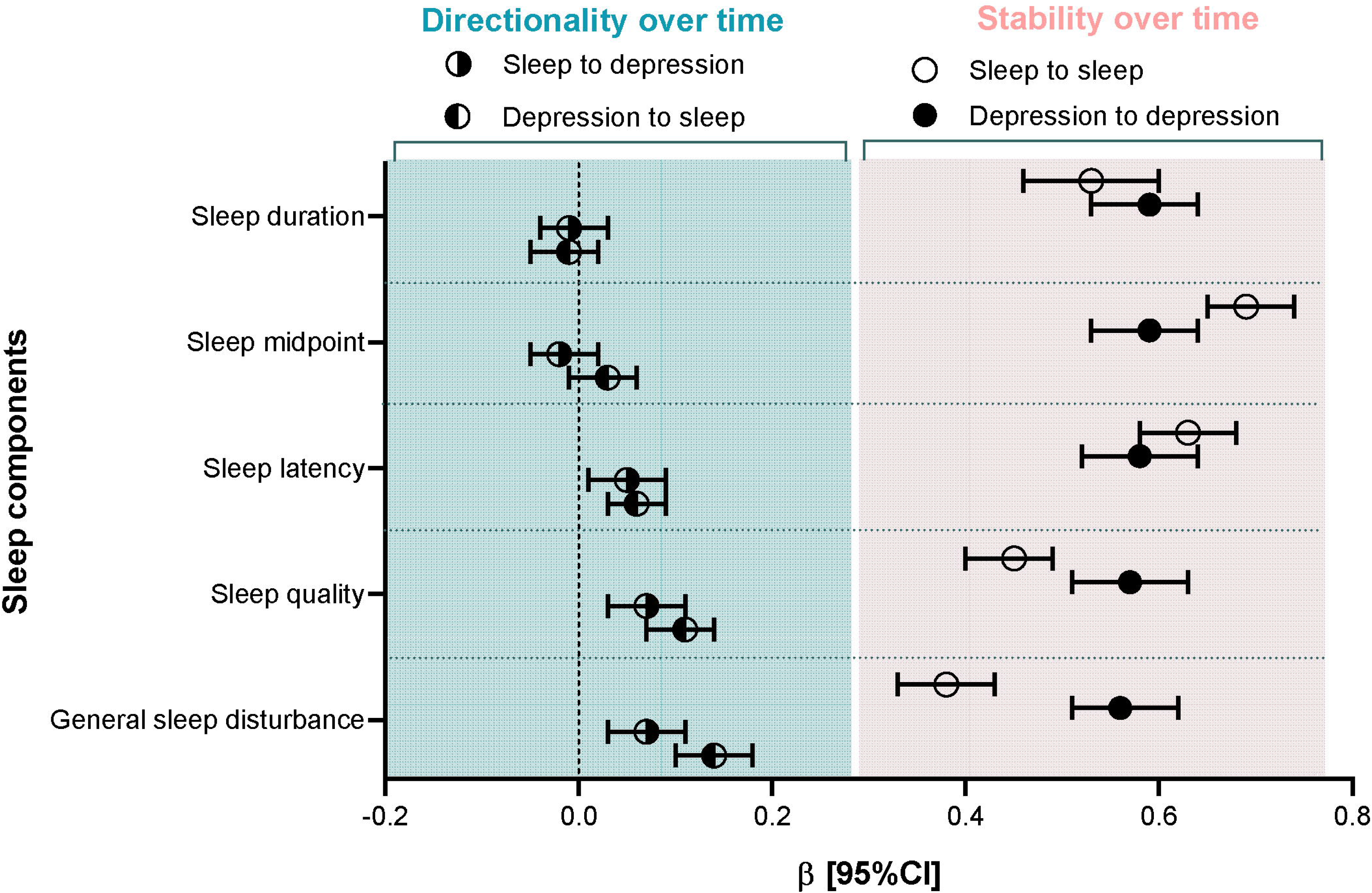
Bidirectional associations between maternal depressive symptoms and sleep with constrained path weights. *Figure note:* This figure shows the standardized regression coefficients (β, reflecting the standard deviation increase in outcome for each standard deviation increase in exposure) and their 95% confidence interval (95%CI) for the bidirectional associations between sleep and depressive symptoms (left, with green background), as well as the stability of each construct over time (right, with pink background), estimated with an Autoregressive Latent Trajectory Model with Structured Residuals (ALT-SR). The weights of all autoregressive paths (stability over time) and all cross-lagged paths (directionality over time) were separately constrained to be equal, given the moderate to high autocorrelations of sleep and depression over time (see heatmap in Figure S2). Model fit: CFI>0.75 and RMSEA <0.06. The model was adjusted for: maternal age at enrollment, country of birth, educational status, household net income monthly, living with a partner [yes/no], parity before enrollment, body mass index at enrollment [kg/m^2^], smoking history in the past 12 months, alcohol use history in the past 12 months, and gestational age at each time point.

When examining stability over time, poor sleep (β_sleepàsleep_=0.38 to 0.69 across sleep domains, all p < 0.001, void circle) and depressive symptoms (β_depressionàdepression_ =0.56 to 0.59 across sleep domains, all p < 0.001, solid circle) were each associated with their respective symptoms at subsequent timepoints. This indicates moderate-to-high stability over time, with one standard deviation (SD) increase at one timepoint associating with a 0.4–0.7 SD increase in the same domain at the next.

When examining the directionality over time, we observed 1-SD increase of depressive symptoms was associated with 0.06-SD increase of subsequent sleep latency (β_depressionàsleep latency_ = 0.06, 95% CI:0.03–0.09, solidàvoid circle), and in the reverse direction 1-SD increase of sleep latency was linked to 0.05-SD increase depressive symptoms (β_sleep latencyàdepression_ = 0.05, 95% CI:0.01–0.09, voidàsolid circle). For sleep quality and general sleep disturbance, associations were slightly stronger from depressive symptoms to sleep (sleep quality:β_depressionàsleep quality_ = 0.11, 95% CI:0.07–0.14; general sleep disturbance:β_depressionàdisturbance_ = 0.14, 95% CI:0.10–0.18, solidàvoid circle) than from sleep to depressive symptoms (β_sleep quality/disturbanceàdepression_ = 0.07 for both, 95% CIs:0.03–0.11, voidàsolid circle). No significant bidirectional associations were found between depressive symptoms and either sleep duration or midpoint (all p > 0.05).

### The role of infant sleep

Infant sleep latency was the only infant sleep component that mediated the association from maternal depressive symptoms to maternal sleep latency (indirect effect: β_depressionàinfant sleep latencyàmaternal sleep latency_=0.28, 95% CI:0.06–0.63, accounting for 9.74% of total effect; Table 2). In contrast, in the reverse path from maternal sleep latency to depressive symptoms, no mediation effect was observed. Infant sleep latency did exert a small-to-moderate moderating effect on the bidirectional association between maternal depressive symptoms and sleep (interaction: β_depression x infant sleep latencyàmaternal sleep latency_=0.11, 95% CI:0.03–0.19, β_maternal sleep latency x infant sleep latencyàdepression_=0.01, 95% CI:0.004–0.02), with a stronger bidirectional association in women with higher levels of infant sleep latency. We did not observe significant mediation or moderation for other infant sleep components in the associations between maternal depressive symptoms and sleep latency, sleep quality, and general sleep disturbance (all p>0.05).

**Table 2:** The mediating and moderating role of infant sleep in the association between maternal depressive symptoms and maternal sleep.

| Paths | Infant frequency of awakenings at night |  | Infant sleep latency |  | Infant nocturnal sleep duration |  |
| --- | --- | --- | --- | --- | --- | --- |
| | $\beta$ [95% CI] | *P* value | $\beta$ [95% CI] | *P* value | $\beta$ [95% CI] | *P* value |
| *Mediation path* |  |  |  |  |  |  |
| Depression -> M -> Sleep latency | 0.02 [-0.04, 0.12] | 0.610 | **0.28 [0.06, 0.63]** | **0.046** | 0.05 [-0.02, 0.18] | 0.313 |
| Depression -> M -> Sleep quality | 0.06 [-0.02, 0.16] | 0.188 | 0.05 [0.01, 0.14] | 0.099 | 0.06 [-0.02, 0.16] | 0.188 |
| Depression -> M -> General sleep disturbance | 0.23 [-0.08, 0.57] | 0.177 | 0.13 [0, 0.35] | 0.135 | 0.15 [0.02, 0.40] | 0.100 |
| Sleep latency -> M -> Depression | <0.001 [-0.001, 0.002] | 0.933 | 0.002 [-0.002, 0.007] | 0.377 | 0.001 [<-0.001, 0.004] | 0.436 |
| Sleep quality -> M -> Depression | 0.003 [-0.001, 0.012] | 0.268 | 0.003 [-0.001, 0.013] | 0.340 | 0.002 [-0.002, 0.01] | 0.506 |
| General sleep disturbance -> M -> Depression | 0.001 [<-0.001, 0.004] | 0.241 | 0.001 [<-0.001, 0.005] | 0.352 | 0.001 [<-0.001, 0.004] | 0.511 |
| *Moderation path* |  |  |  |  |  |  |
| Depression*M -> Sleep latency | 0.56 [-0.31, 1.42] | 0.207 | **0.11 [0.03, 0.19]** | **0.008** | -0.001 [-0.02, 0.01] | 0.237 |
| Depression*M -> Sleep quality | 0.13 [-0.21, 0.47] | 0.445 | 0.02 [-0.02, 0.05] | 0.308 | 0.001 [-0.004, 0.01] | 0.656 |
| Depression*M -> General sleep disturbance | 0.52 [-0.57, 1.60] | 0.351 | 0.06 [-0.04, 0.16] | 0.225 | -0.001 [-0.02, 0.02] | 0.870 |
| Sleep latency*M -> Depression | 0.04 [-0.17, 0.26] | 0.685 | **0.01 [0.004, 0.02]** | **0.001** | -0.003 [-0.007, 0.001] | 0.143 |
| Sleep quality*M -> Depression | -0.03 [-0.63, 0.56] | 0.916 | 0.03 [-0.004, 0.07] | 0.080 | - | - |
| General sleep disturbance*M -> Depression | 0.16 [-0.27, 0.58] | 0.467 | 0.03 [-0.004, 0.06] | 0.089 | -0.001 [-0.008, 0.007] | 0.864 |

*Table note:* This table presents the mediating and moderating roles of infant sleep within the structural equation model framework, based on 1,000 bootstrap iterations. Analyses were restricted to domains with significant bidirectional and autoregressive effects (sleep latency, sleep quality, and general sleep disturbance). Standardized regression coefficients ( $\beta$ ; reflecting the standard deviation increase in outcome for each standard deviation increase in exposure), 95% confidence intervals (CI), and infant sleep parameters as mediators or moderators (M; frequency of night awakenings, sleep latency, nocturnal sleep duration) are reported. Infant sleep latency mediated 9.74% of effect of depression on maternal sleep latency and exerted a small-to-moderate moderating effect on the bidirectional association between maternal depression and sleep latency. Covariates were adjusted in the model, and included maternal age at enrollment, country of birth, educational status, household net income monthly, living with a partner (yes vs. no), parity before enrollment (nulliparous vs. primiparous or multiparous), body mass index at enrollment (kg/m<sup>2</sup>), smoking (never vs. ever or current), alcohol use in the past 12 months (yes vs. no), and gestational age at each time point. Bold values indicate $p < 0.05$ . Dashes (–) indicate paths not estimated or not applicable.

### Sensitivity analysis

Main findings remained consistent when we allowed path estimates to vary across timepoints (Supplementary Figure S3, Model fit CFI>0.75 and RMSEA<0.06). Both poor sleep and depressive symptoms were moderately-to-strongly associated with their respective symptoms at subsequent timepoints (all p<0.05). There were no bidirectional associations between perinatal sleep components and depressive symptoms from preconception to the first trimester (all p>0.05). Depressive symptoms in the first trimester were associated with longer *sleep latency* in the third trimester (β_depression 1st trimesteràsleep latency 3rd trimester_= 0.07, 95% CI:0.02–0.12), but not in the reverse direction. From the third trimester to six months postpartum, sleep latency was associated with increased depressive symptoms at six months postpartum (β = 0.09 _sleep latency 3rd trimesteràdepression 6m postpartum_, 95% CI:0.002–0.19), but not in the reverse direction. For *sleep quality*, higher depressive symptoms scores in the first (β_depression 1st trimesteràsleep quality 3rd trimester_ = 0.10, 95% CI:0.05–0.15) and third trimester (β_depression 3rd trimesteràsleep quality 6m postpartum_= 0.14, 95% CI:0.07–0.21) were associated with poorer sleep quality at later timepoints. In the reverse path, sleep quality associated with later depressive symptoms in the first trimester (β_sleep quality 1st trimesteràdepression 3rd trimester_= 0.07, 95% CI:0.001–0.13) as well as in the third trimester (β_sleep quality 3rd trimesteràdepression 6m postpartum_= 0.14, 95% CI:0.07–0.20). For *general sleep disturbance*, higher depressive symptom scores associated with more disturbance both in the first (β_depression 1st trimesteràdisturbance 3rd trimester_ = 0.19, 95% CI:0.14–0.24) and third trimesters (β_depression 3rd trimesteràdisturbance 6m postpartum_ = 0.15, 95% CI:0.08–0.22), as well as in the reverse direction (first trimester: β^disturbance 1st trimesteràdepression 3^_rd trimester_ = 0.09, 95% CI:0.02–0.15; third trimester: β_disturbance 3rd trimesteràdepression 6m postpartum_= 0.11, 95% CI:0.03–0.19).

Results were consistent after excluding the overlapping EPDS item (results not shown) in main analyses with constrained weights.

## Discussion

In this population-based cohort study, we investigated the bidirectional association between maternal depressive symptoms and multiple sleep components from preconception through 12 months postpartum. We identified significant bidirectional associations of depressive symptoms with sleep latency, sleep quality, and general sleep disturbance, whereas no such associations were found for sleep duration or sleep midpoint. Notably, infant sleep latency emerged as the sole infant sleep component acting as both a mediator and moderator in the association between maternal depressive symptoms and maternal sleep latency. Although effect sizes were modest, the consistency of findings across sensitivity analyses underscores the robustness of these developmental dynamics.

Our findings extend previous research that has largely focused on the unidirectional pathway from prenatal sleep to subsequent postnatal depressive symptoms (Emamian et al., 2019; Fu et al., 2023; Lawson et al., 2015; Li et al., 2023; Maghami et al., 2021; Sobol et al., 2023). While a few studies have explored bidirectionality starting in the postpartum period, they have predominantly identified unidirectional effects—specifically, that insomnia and poor sleep quality predict subsequent depression, but not vice versa (Astbury et al., 2025; Okun et al., 2025). By expanding the observation window to include preconception and extending it to 12 months postpartum, our study captured bidirectional dynamics that may have been missed in studies with narrower timeframes. Our sensitivity analyses revealed that the pathway from depressive symptoms to sleep became more pronounced from the first trimester onwards. This pattern may reflect a cumulative developmental effect, where the physiological and psychological burden of pregnancy and postpartum care progressively strengthens the impact of maternal mood on sleep regulation, but also may be due to the smaller sample size at earlier timepoints, which may have limited the sensitivity to detect bidirectional effects at preconception already. In addition, studying the association between sleep and depressive symptoms is complex, as poor sleep is often considered a symptom of depression (American Psychiatric Association, 2013), and consistent with this, sleep-related items are embedded in many depressive symptoms questionnaires. However, bidirectional associations remained robust in sensitivity analyses excluding overlapping items, indicating that the observed effects were not driven by measurement overlap.

Importantly, the bidirectional nature of these associations varied by sleep component. Significant effects were evident for sleep latency, quality, and disturbance, but not for duration or midpoint. The pathways from depressive symptoms to sleep quality and disturbance appeared slightly stronger than the reverse (though confidence intervals overlapped), while effects for sleep latency were comparable in both directions. This pattern may reflect differences between sleep components. Sleep quality and general disturbance are inherently subjective and may be more susceptible to the cognitive and emotional distortions characteristic of depression, such as negative appraisal and rumination (Moulds et al., 2022). In contrast, sleep latency represents a distinct behavioral challenge (Kalmbach et al., 2021). These findings highlight the multidimensionality of sleep and the necessity of analyzing components separately to uncover specific intervention targets.

Shared etiological mechanisms likely underpin these bidirectional associations. Across the life course, sleep and depression share biological pathways, including genetic vulnerability (Hamilton et al., 2023), hypothalamic–pituitary–adrenal (HPA) axis dysregulation (Asarnow, 2020), and circadian rhythm dysregulation through the suprachiasmatic nucleus (Vadnie and McClung, 2017). In the perinatal context, these are compounded by dramatic hormonal fluctuations (Okun, 2016), as well as psychological processes. For example, pregnancy-related hyperarousal and anxiety can prolong sleep latency or distort subjective sleep evaluations, while simultaneously increasing vulnerability to depressive symptoms (Kalmbach et al., 2021; Moulds et al., 2022). Although not directly assessed, these pathways provide a plausible framework for the reinforcing cycle observed in our data.

A key novelty of this study is the identification of infant sleep latency as a dual mechanism. As a mediator, infant sleep latency explained approximately 10% of the effect of maternal depressive symptoms on maternal sleep latency. Potentially, elevated maternal depressive symptoms may influence biological pathways (e.g., serotonin dysregulation, altered EEG patterns) and caregiving routines due to parenting stress, leading to difficulties in settling the infant, which in turn prolongs the mother’s own time to fall asleep (Galbally et al., 2009; Grieve et al., 2019; Morales-Muñoz et al., 2018). Consistent with this interpretation, longitudinal research has shown that higher maternal sensitivity in early childhood associated with fewer child sleep problems years later, highlighting the long-term role of sensitive caregiving in promoting healthy sleep regulation (Middlemiss et al., 2017). As a moderator, longer infant sleep latency strengthened the bidirectional association between maternal depressive symptoms and maternal sleep latency. This suggests that a “difficult-to-settle” infant creates a high-risk context that amplifies maternal vulnerability. The specificity of this finding to sleep latency, rather than any other sleep component is striking. We propose this is driven by temporal alignment: both maternal and infant sleep initiation occur simultaneously at the start of the night. This synchronization creates a period of intense behavioural and emotional interdependence. An infant who struggles to fall asleep directly delays the mother’s sleep onset and likely heightens her pre-sleep arousal and anxiety. Shared genetic influences on sleep may further contribute to this alignment (Lane et al., 2023). In contrast, infant sleep duration or night awakenings span the entire night and may be less tightly synchronized in real-time. Although reporter bias cannot be excluded, it is unlikely to entirely explain the specificity, as other infant sleep outcomes were also mother reported. Overall, our study suggests that infant sleep latency may serve as a valuable intervention target for postpartum intervention, especially when preventive opportunities during preconception or pregnancy have been missed.

Our study has several strengths. First, the use of a large, population-based cohort with repeated assessments from preconception through one year postpartum provided an observation window across various important developmental transitions. Second, we employed comprehensive measurements of multiple sleep components in both mothers and infants, enabling a multidimensional perspective on perinatal sleep. Third, we simultaneously examined the role of infant sleep as both a mediator and a moderator to offer novel insight into its role in the bidirectional association between maternal sleep and depressive symptoms.

Several limitations should be considered when interpreting the results. First, participants were predominantly of higher socioeconomic status, which may limit the generalizability of our results to populations with lower socioeconomic status. Second, both maternal and infant sleep were measured via mother-reported questionnaires. Recall bias may occur if mothers misremember their own or their infant’s sleep patterns. Common method bias may also arise because the same reporter was used for both maternal and infant sleep, inflating associations between them. Future studies should employ objective sleep measures, such as actigraphy, to complement our findings. Third, we used two different measurements of maternal depressive symptoms across timepoints. While EPDS has demonstrated validity beyond the perinatal context and our study showed they are highly correlated over time, it remains unknown whether these two scales are comparable within the same sample (Matijasevich et al., 2014). Future studies should test if ASR and EPDS capture depressive symptoms similarly across time. Fourth, missing data frequencies in this study were highest in the preconception period, as women could initiate study participation either before conception or during pregnancy. This inclusion strategy inherently resulted in two slightly different subsamples, with some women lacking preconception information. To address this, we applied full information maximum likelihood algorithms in analyses, which allows the inclusion of all participants by estimating parameters based on available data under the assumption of missing at random. Although this approach makes full use of our sample, the imbalance in available preconception data should be considered when interpreting the results. Fifth, some residual confounding may exist due to unmeasured covariates, such as occupational status across pregnancy.

## Conclusion

This study provides robust evidence for bidirectional associations between depressive symptoms and sleep latency, quality, and disturbance, starting as early as preconception. Postpartum infant sleep latency further acted as both a mediator and moderator of the sleep latency-depressive symptoms link. Future research should replicate these findings using objective sleep measures and in more socioeconomically and culturally diverse populations. Integrating coordinated support for maternal sleep and mental health into preconception and antenatal care, alongside infant sleep guidance in postnatal care, may provide modifiable leverage points to redirect the bidirectional sleep-depressive symptoms association.

## Acknowledgements

We acknowledge the following funding scources. The general design of Generation R Next Study is made possible by financial support from the Erasmus Medical Center and the Erasmus University Rotterdam, the Netherlands Organization for Health Research and Development (ZonMW), the Netherlands Organisation for Scientific Research (NWO), the Ministry of Health, Welfare and Sport and the Ministry of Youth and Families. This project is a part of HappyMums, funded by European Union (Grant Agreement No.101057390). FM received a scholarship from China Scholarships Council (CSC, 202206220040). HM was supported by Stichting Volksbond Rotterdam, the Netherlands Organization for Health Research and Development [Aspasia grant No.015.016.056].

The study sponsors had no role in the study design, collection, analysis and interpretation of data, writing of the report, or in the decision to submit the paper for publication.

## Data availability

Data sharing Generation R: The analysis plan, statistical code, and data from this study are available upon reasonable request to the director of the Generation R Study, subject to local, national, and European rules and regulations.

## Ethical Approval Statement

The study procedures were approved by the Medical Ethical Committee of the Erasmus Medical Center (MEC-2016-589, December 2016). Written informed consent was obtained from all participants.

