## Supplement tables and figures for "From preconception to postpartum: bidirectional associations between sleep and depression and the role of infant sleep in a population-based cohort"

### Supplementary materials

#### Content

**Supplementary Tables**

Table S1: Non-response analysis

Table S2: The association between sleep and depressive symptoms from preconception to postpartum when constraining path weights

**Supplementary Figures**

Figure S1: Distribution of data availability over the study period (n = 2,294)

Figure S2: Heatmap of temporal correlations within maternal depressive symptoms and sleep components from preconception to 12 months postpartum

Figure S3: The association between sleep and depressive symptoms with varying path weights

#### Supplementary Tables

Table S1: Non-response analysis

| Demographic variables | Study population (n=2,294), Mean ± SD / No. (%) | Excluded population * (n=469), Mean ± SD / No. (%) | P value |
|---|---|---|---|
| Age at enrollment, yrs. | 31.7 ± 4.2 | 29.4 ± 5.4 | <0.001 |
| Country of birth | | | <0.001 |
| the Netherlands | 1423 (63.6%) | 91 (34.1%) | |
| Outside the Netherlands | 814 (36.4%) | 176 (65.9%) | |
| Education status | | | <0.001 |
| Primary or secondary [vocational] | 603 (27.2%) | 156 (59.8%) | |
| Bachelor's or associate degree | 681 (30.7%) | 55 (21.1%) | |
| Master's degree | 933 (42.1%) | 50 (19.2%) | |
| Household net income, monthly | | | <0.001 |
| Less than €3000 | 458 (22.4%) | 132 (57.1%) | |
| Equal or more than €3000 | 1583 (77.6%) | 99 (42.9%) | |
| Living with partner | | | <0.001 |
| Yes | 1903 (87.2%) | 106 (71.1%) | |
| No or Other | 280 (12.8%) | 43 (28.9%) | |
| Parity before enrollment | | | <0.001 |
| Nulliparous | 1450 (65.8%) | 73 (45.1%) | |
| Primiparous or multiparous | 753 (34.2%) | 89 (54.9%) | |
| Body mass index at enrollment, kg/m$^2$ | 24.2 ± 4.4 | 25.7 ± 5.3 | <0.001 |
| Smoking status in the past 12 months | | | <0.001 |
| Never | 1164 (55.5%) | 81 (46.8%) | |
| Any | 932 (44.5%) | 92 (53.2%) | |
| Alcohol use status in the past 12 months | | | <0.001 |
| Never | 489 (21.9%) | 70 (38.0%) | |
| Ever or current | 1746 (78.1%) | 114 (62.0%) | |
| Child sex | | | 1 |
| Male | 1188 (52.7%) | 242 (52.7%) | |
| Female | 1068 (47.3%) | 217 (47.3%) | |

*Table Note:* * Women with no data on sleep (n = 441) or depressive symptoms (n = 28) were excluded (n=469 in total). The percentages of missing values in excluded populations across all variables ranged from 0.1 to 68.2%, with the highest percentage of missing found in living with partner. The percentages of missing in included populations across all variables ranged from 0.1 to 11.0%, with the highest percentage of missing in maternal net family income monthly.

Table S2: The association between sleep and depressive symptoms from preconception to postpartum when constraining path weights

| **Path** | **Sleep duration**<br>**β [95% CI]** | **Sleep midpoint**<br>**β [95% CI]** | **Sleep latency**<br>**β [95% CI]** | **Sleep quality**<br>**β [95% CI]** | **General sleep disturbance**<br>**β [95% CI]** |
|---|---|---|---|---|---|
| *Stability over time* | | | | | |
| Depression -> Depression | **0.59 [0.53, 0.64]** | **0.59 [0.53, 0.64]** | **0.58 [0.52, 0.64]** | **0.57 [0.51, 0.63]** | **0.56 [0.51, 0.62]** |
| Sleep -> Sleep | **0.53 [0.46, 0.60]** | **0.69 [0.65, 0.74]** | **0.63 [0.58, 0.68]** | **0.45 [0.40, 0.49]** | **0.38 [0.33, 0.43]** |
| *Directionality over time* | | | | | |
| Depression -> Sleep | -0.01 [-0.05, 0.02] | 0.03 [-0.01, 0.06] | **0.06 [0.03, 0.09]** | **0.11 [0.07, 0.14]** | **0.14 [0.10, 0.18]** |
| Sleep -> Depression | -0.01 [-0.04, 0.03] | -0.02 [-0.05, 0.02] | **0.05 [0.01, 0.09]** | **0.07 [0.03, 0.11]** | **0.07 [0.03, 0.11]** |
| *Concurrent associations* | | | | | |
| Depression at T1 -> Sleep at T1 | -0.06 [-0.14, 0.02] | **0.15 [0.05, 0.25]** | **0.18 [0.09, 0.27]** | **0.81 [0.74, 0.88]** | **0.27 [0.19, 0.36]** |
| Depression at T2 -> Sleep at T2 | -0.03 [-0.07, 0.01] | 0.015 [-0.01, 0.04] | **0.05 [0.01, 0.09]** | **0.18 [0.15, 0.22]** | **0.17 [0.13, 0.21]** |
| Depression at T3 -> Sleep at T3 | -0.03 [-0.07, 0.01] | 0.015 [-0.01, 0.04] | **0.05 [0.01, 0.09]** | **0.18 [0.15, 0.22]** | **0.17 [0.13, 0.21]** |
| Depression at T5 -> Sleep at T5 | -0.03 [-0.07, 0.01] | 0.015 [-0.01, 0.04] | **0.05 [0.01, 0.09]** | **0.18 [0.15, 0.22]** | **0.17 [0.13, 0.21]** |

*Table note:* This table shows the standardized regression coefficients (β; reflecting the standard deviation increase in outcome for each standard deviation increase in exposure) and their 95% confidence interval (95%CI) for the bidirectional associations between sleep and depressive symptoms, as well as the stability of each construct over time, estimated with an Autoregressive Latent Trajectory Model with Structured Residuals (ALT-SR).

The weights of all autoregressive paths (stability over time) and all cross-lagged paths (directionality over time) were separately constrained to be equal, given the moderate to high autocorrelations of sleep and depressive symptoms over time (see heatmap in Figure S2).

The models were adjusted for: maternal age at enrollment, country of birth, educational status, household net income monthly, living with a partner (yes vs. no), parity before enrollment (nulliparous vs. primiparous or multiparous), body mass index at enrollment ($kg/m^2$), smoking (never vs. ever or current), alcohol use in the past 12 months (yes vs. no), and gestational age at each time point. Model fit: CFI >0.75 and RMSEA <0.06

Bold text indicates significant findings with p-value <0.05.

#### Supplementary Figures

Figure S1: Distribution of data availability over the study period (n = 2,294)

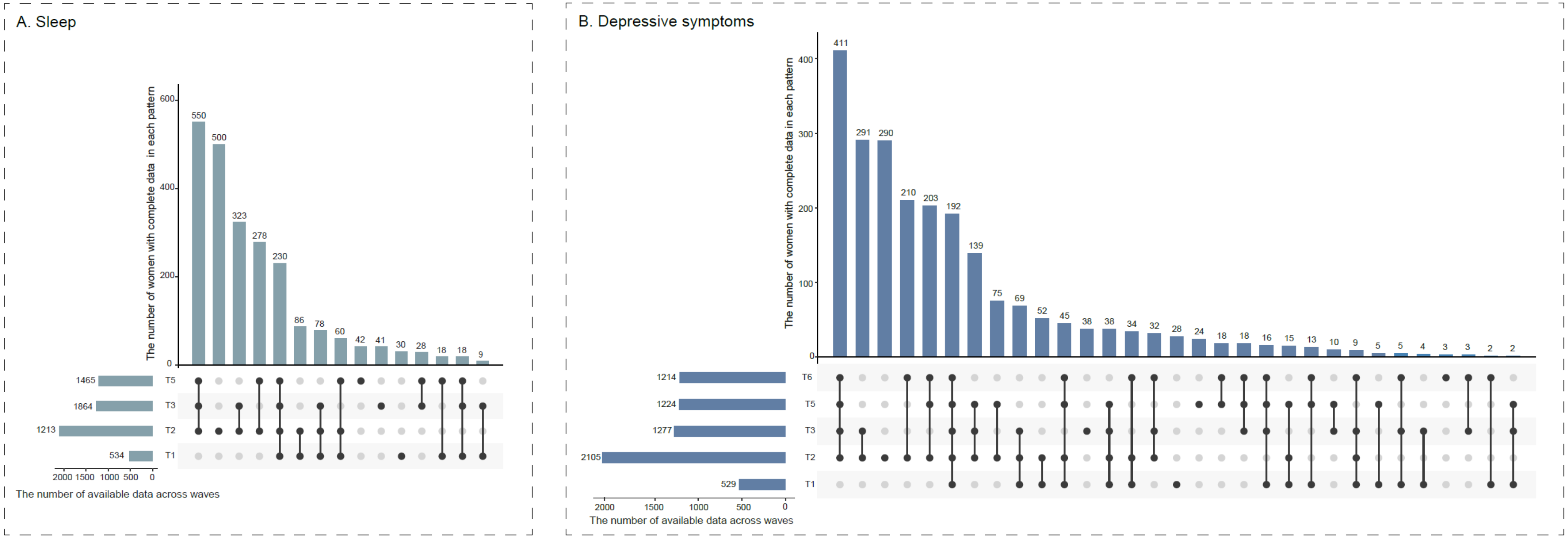


*Figure note:* This figure presents the distribution of data availability over time for sleep (Panel A, left) and depressive symptoms (Panel B, right). The line plots at the bottom depict the combination of data availability, with points connected to indicate the specific timepoints at which complete data are available. The vertical bar charts at the top represent the number of cases for each combination pattern, while the horizontal bar charts at the left-bottom show the accumulated number of cases at each timepoint. Timepoints are defined as follows: T1 = preconception, T2 = 1st trimester, T3 = 3rd trimester, and T5 = six months postpartum, T6 = 12 months postpartum.

Figure S2: Heatmap of temporal correlations within maternal depressive symptoms and sleep components from preconception to 12 months postpartum

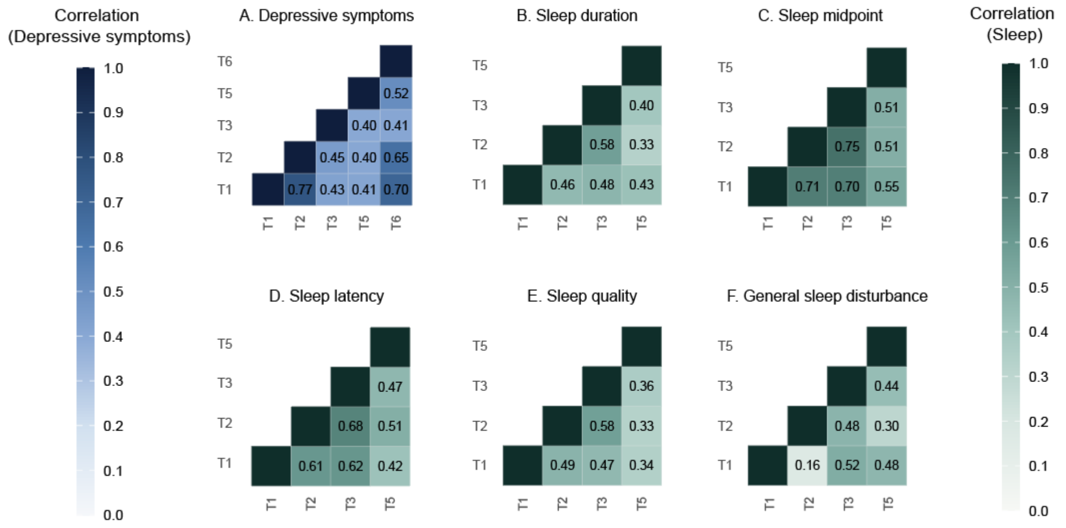


*Figure note:* This figure shows the bivariate correlation (Pearson r) within depressive symptoms (blue) and each sleep component (green) and over time. Timepoints are defined as follows: T1 = preconception, T2 = first trimester, T3 = third trimester, and T5 = six months postpartum, T6 = 12 months postpartum. Depressive symptoms at T1, T2 and T6 were measured by the Adult Self Report subscale and depressive symptoms at T3 and T5 were measured by the Edinburgh Postnatal Depression Scale. T4 representing the first month postpartum, when infant sleep was measured, is not shown in this plot.

Figure S3: The association between sleep and depressive symptoms with varying path weights

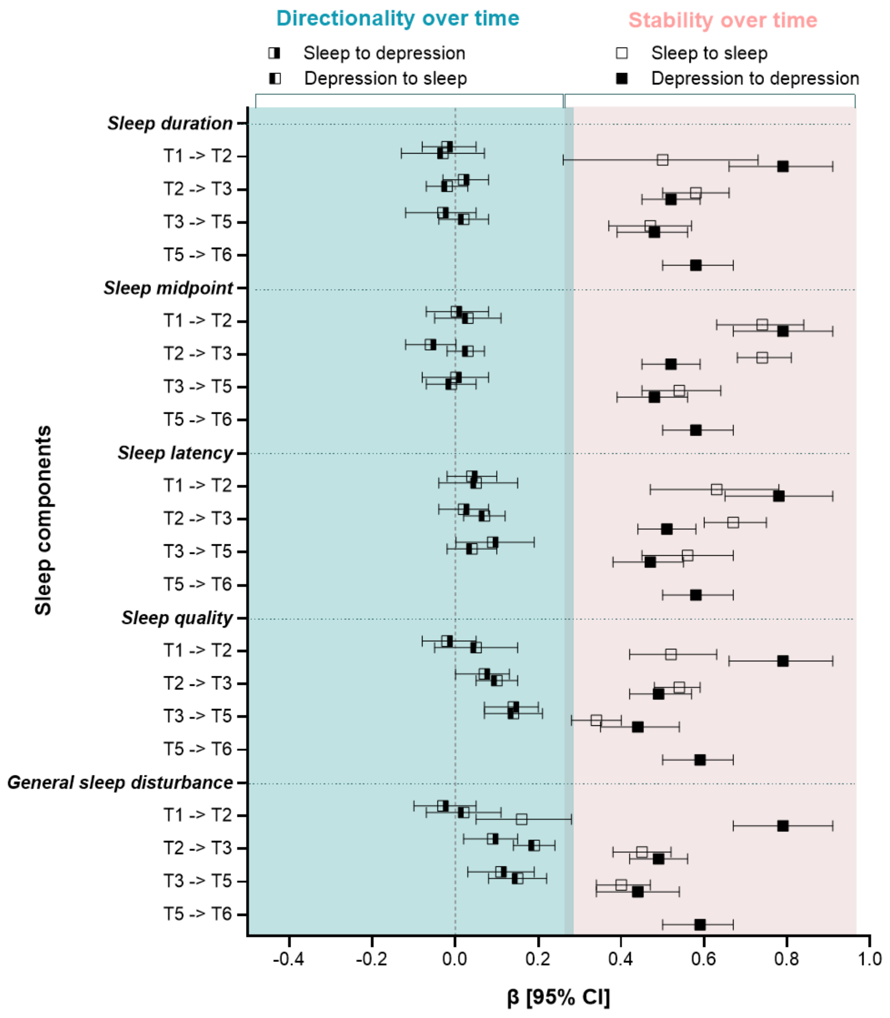


*Figure note:* This figure shows the standardized regression coefficients (β, reflecting the standard deviation increase in outcome for each standard deviation increase in exposure) and their 95% confidence interval (95%CI) for the bidirectional associations between sleep and depressive symptoms (left, with green background), as well as the stability of each construct over time (right, with pink background), estimated with an Autoregressive Latent Trajectory Model with Structured Residuals (ALT-SR). All autoregressive and cross-lagged paths were freely estimated (i.e. not constrained to be equal across timepoints). Model fit: CFI >0.75 and RMSEA <0.06.

Timepoints are defined as follows: T1 = preconception, T2 = first trimester, T3 = third trimester, and T5 = six months postpartum, T6 = 12 months postpartum. T4 representing first months postpartum when infant sleep was measured are not shown in this plot. The model was adjusted for: maternal age at enrollment, country of birth, educational status, household net income monthly, living with a partner (yes vs. no), parity before enrollment (nulliparous vs. primiparous or multiparous), body mass index at enrollment (kg/m$^2$), smoking (never vs. ever or current), alcohol use in the past 12 months (yes vs. no), and gestational age at each time point
